# Protection afforded by the BNT162b2 and mRNA-1273 COVID-19 vaccines in fully vaccinated cohorts with and without prior infection

**DOI:** 10.1101/2021.07.25.21261093

**Authors:** Laith J. Abu-Raddad, Hiam Chemaitelly, Houssein H. Ayoub, Hadi M. Yassine, Fatiha M. Benslimane, Hebah A. Al Khatib, Patrick Tang, Mohammad R. Hasan, Peter Coyle, Zaina Al Kanaani, Einas Al Kuwari, Andrew Jeremijenko, Anvar Hassan Kaleeckal, Ali Nizar Latif, Riyazuddin Mohammad Shaik, Hanan F. Abdul Rahim, Gheyath K. Nasrallah, Mohamed Ghaith Al Kuwari, Adeel A. Butt, Hamad Eid Al Romaihi, Mohamed H. Al-Thani, Abdullatif Al Khal, Roberto Bertollini

## Abstract

Effect of prior SARS-CoV-2 infection on vaccine protection remains poorly understood. Here, we investigated whether persons vaccinated after a prior infection have better protection against future infection than those vaccinated without prior infection. Effect of prior infection was assessed in Qatar’s population, where the Alpha (B.1.1.7) and Beta (B.1.351) variants dominate incidence, using two national retrospective, matched-cohort studies, one for the BNT162b2 (Pfizer-BioNTech) vaccine, and one for the mRNA-1273 (Moderna) vaccine. Incidence rates of infection among BNT162b2-vaccinated persons, with and without prior infection, were estimated, respectively, at 1.66 (95% CI: 1.26-2.18) and 11.02 (95% CI: 9.90-12.26) per 10,000 person-weeks. The incidence rate ratio was 0.15 (95% CI: 0.11-0.20). Analogous incidence rates among mRNA-1273-vaccinated persons were estimated at 1.55 (95% CI: 0.86-2.80) and 1.83 (95% CI: 1.07-3.16) per 10,000 person-weeks. The incidence rate ratio was 0.85 (95% CI: 0.34-2.05). Prior infection enhanced protection of those BNT162b2-vaccinated, but not those mRNA-1273-vaccinated. These findings may have implications for dosing, interval between doses, and potential need for booster vaccination.

## Main text

Effect of prior acute respiratory syndrome coronavirus 2 (SARS-CoV-2) infection on vaccine protection against acquisition of infection remains poorly understood^1-3^. Qatar launched Coronavirus Disease 2019 (COVID-19) immunization in December 21, 2020, first using the BNT162b2^4^ (Pfizer-BioNTech) vaccine and subsequently adding the mRNA-1273^5^ (Moderna) vaccine^6,7^. As vaccination was scaled up following the FDA-approved protocol, the country experienced two back-to-back SARS-CoV-2 waves from January-June, 2021, which were dominated by the Alpha^8^ (B.1.1.7) and Beta^8^ (B.1.351) variants^6,7,9-11^ (Methods). This provided an opportunity to assess whether persons vaccinated after a prior SARS-CoV-2 infection have better protection against future infection than those vaccinated without prior infection.

Leveraging the national, federated databases that have captured all SARS-CoV-2 vaccinations and PCR testing since the epidemic onset (Methods), we investigated this question using two retrospective, matched-cohort studies. We compared incidence of documented SARS-CoV-2 infection in the national cohort of individuals who completed ≥14 days after the second BNT162b2 vaccine dose, but who had experienced a prior PCR-confirmed infection, with incidence among individuals who completed ≥14 days after the second BNT162b2 dose, but who had not experienced a prior infection, between December 21, 2020-June 6, 2021 (Figure 1). The same comparison was made for the mRNA-1273 vaccine (Figure 2). Cohorts were matched in a 1:1 ratio by sex, 5-year age group, nationality, and calendar week of the first vaccine dose, to control for differences in exposure risk^12,13^ and variant exposure^6,7,9-11^. Reporting of the study followed the STROBE guidelines (Supplementary Table 1).

**Figure 1.**
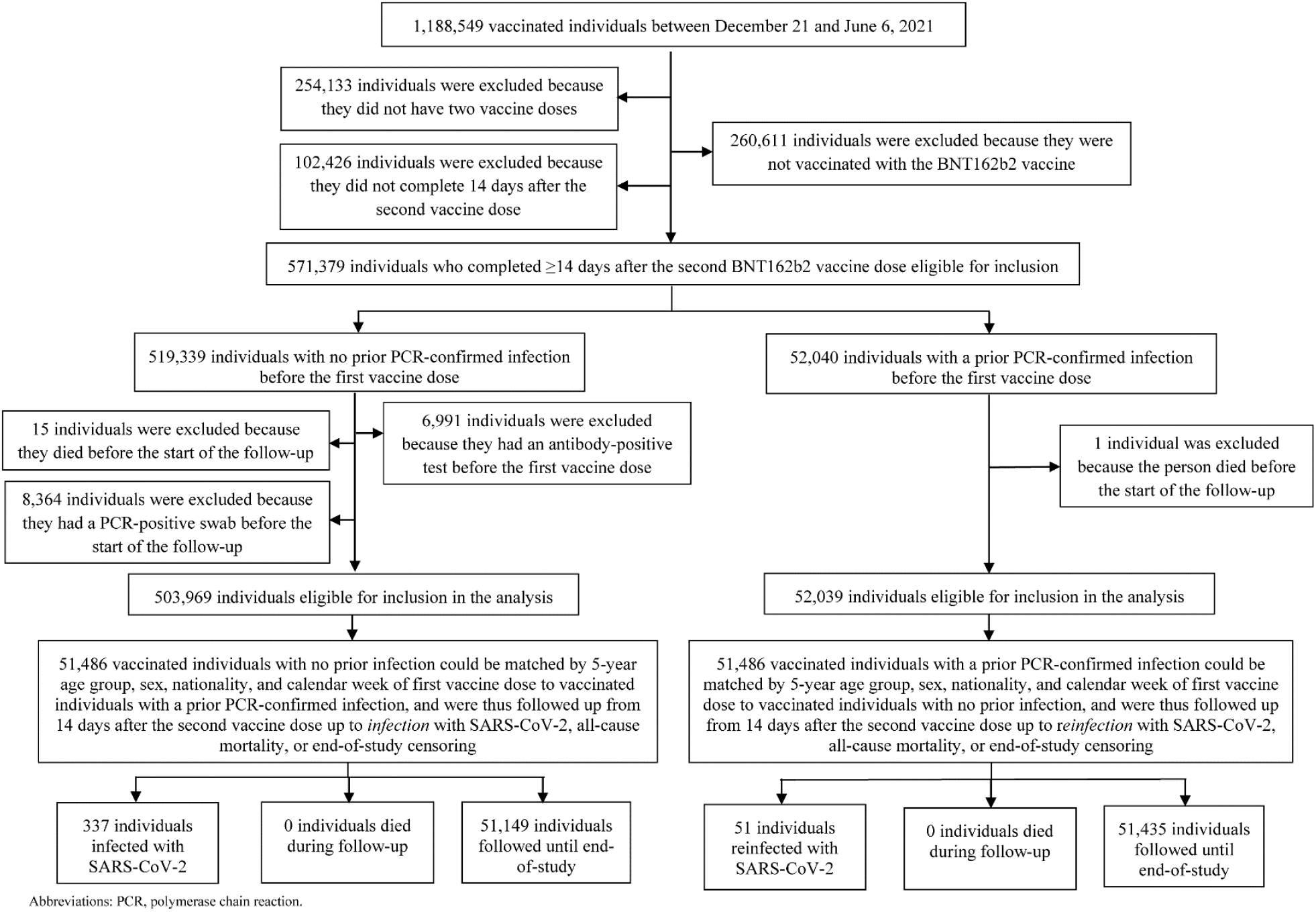
**The process for identifying SARS-CoV-2 infections in the national cohort of individuals who completed ≥14 days after the second BNT162b2 vaccine dose and who had experienced a PCR-confirmed infection before the first dose, compared with the process for identifying SARS-CoV-2 infections in the national cohort of individuals who completed ≥14 days after the second BNT162b2 vaccine dose, but who had experienced no PCR-confirmed infection before the first dose. Cohorts were matched in a 1:1 ratio by sex, 5-year age group, nationality, and calendar week of the first vaccine dose. Total follow-up time among BNT162b2-vaccinated persons, with and without prior infection, was 308,086.0 and 305,891.9 person-weeks, respectively.**

**Figure 2.**
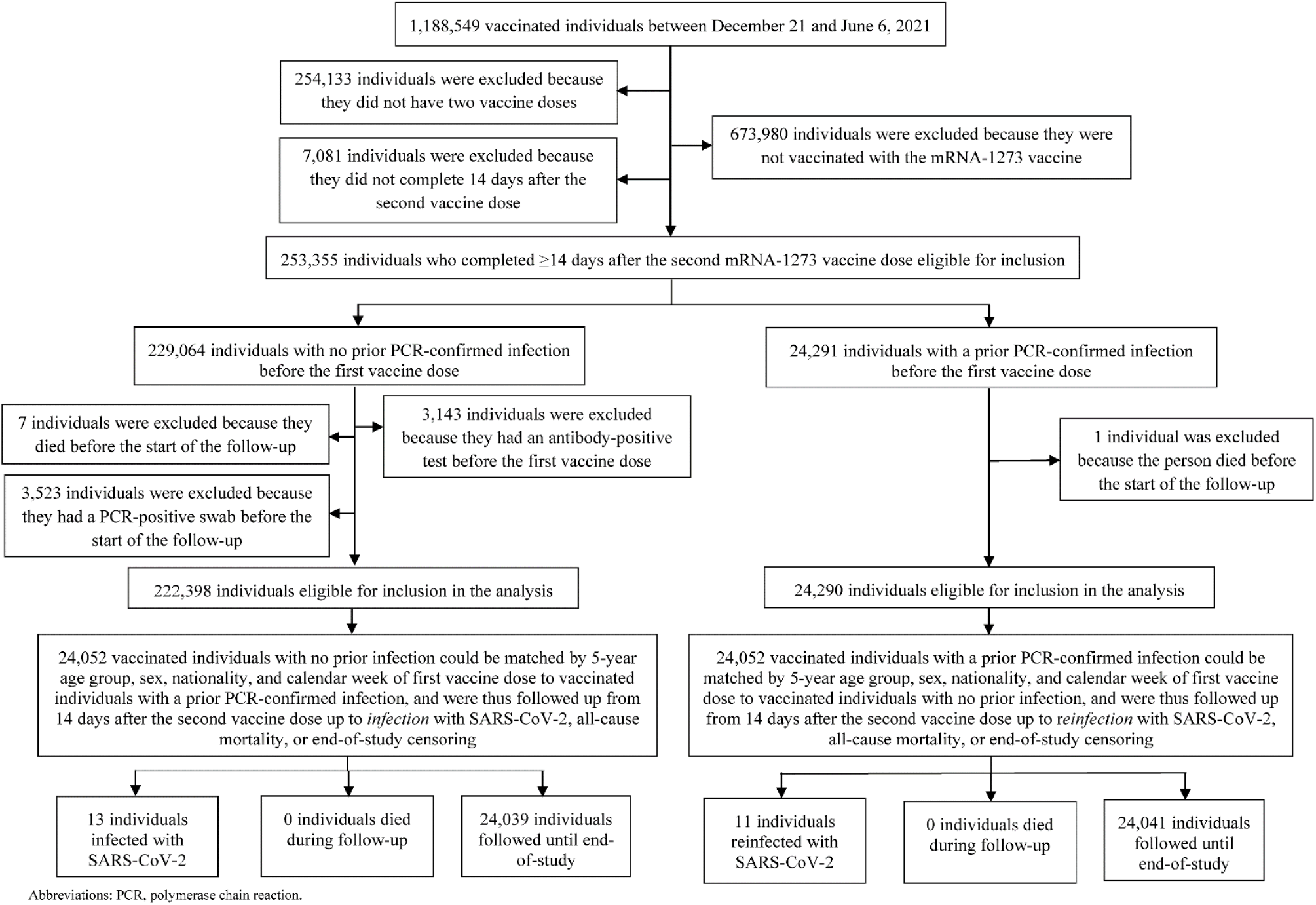
**The process for identifying SARS-CoV-2 infections in the national cohort of individuals who completed ≥14 days after the second mRNA-1273 vaccine dose and who had experienced a PCR-confirmed infection before the first dose, compared with the process for identifying SARS-CoV-2 infections in the national cohort of individuals who completed ≥14 days after the second mRNA-1273 vaccine dose, but who had experienced no PCR-confirmed infection before the first dose. Cohorts were matched in a 1:1 ratio by sex, 5-year age group, nationality, and calendar week of the first vaccine dose. Total follow-up time among mRNA-1273-vaccinated persons, with and without prior infection, was 70,729.9 and 70,872 person-weeks, respectively.**

**Figure 3.**
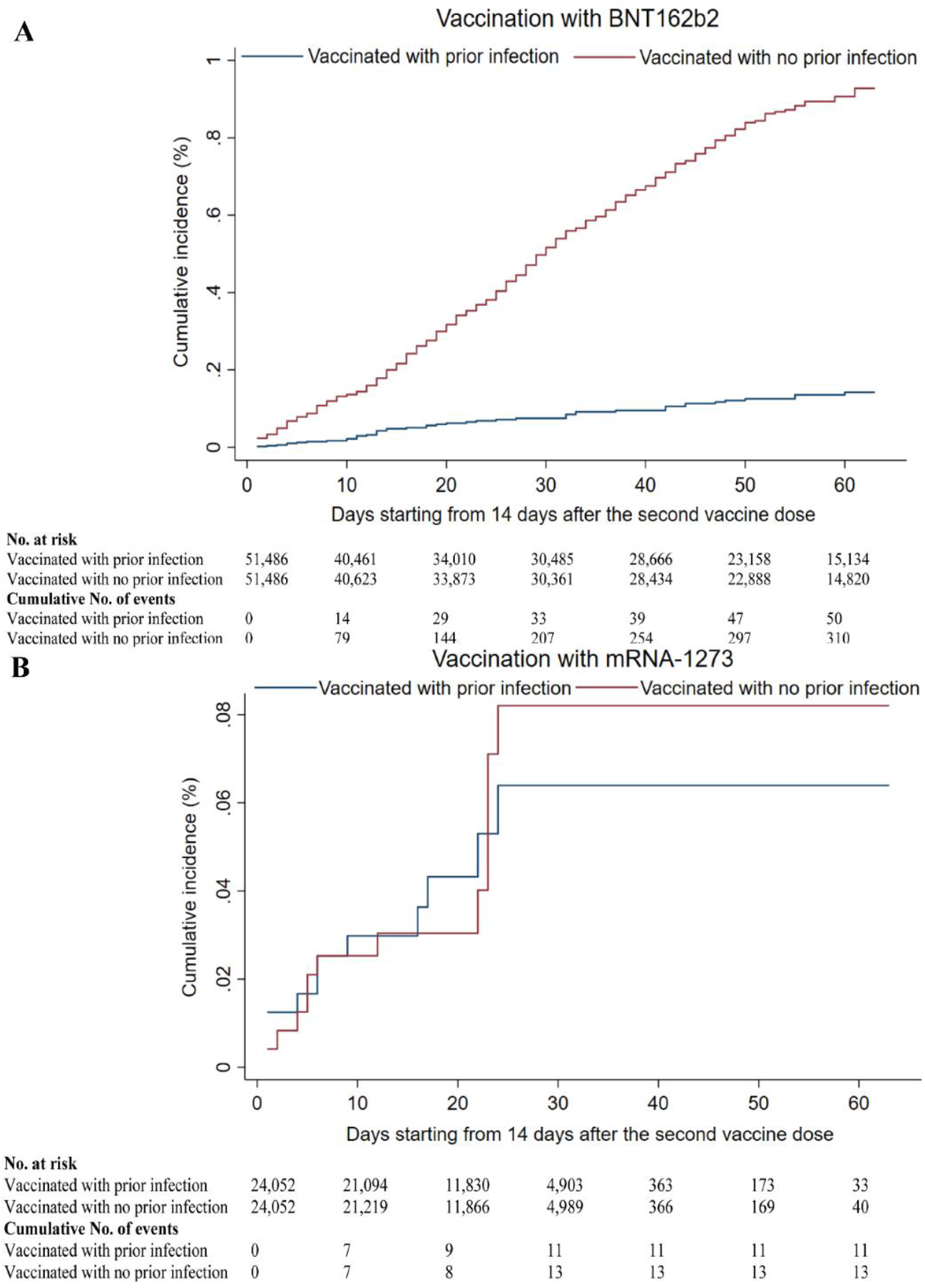
**Kaplan-Meier curves showing the cumulative incidence of documented SARS-CoV-2 infection in the national cohort of individuals who completed ≥14 days after the second vaccine dose and who had a prior PCR-confirmed infection, compared to the cumulative incidence of documented SARS-CoV-2 infection in the matched national cohort of individuals who completed ≥14 days after the second vaccine dose, but without prior PCR-confirmed infection. The curves compare vaccination with A) the BNT162b2 (Pfizer-BioNTech) vaccine and B) the mRNA-1273 vaccine. Cohorts were matched in a 1:1 ratio by sex, 5-year age group, nationality, and calendar week of the first vaccine dose. The curves for a longer time of follow up for only the BNT162b2 vaccine are in Supplementary Figure 1. Vaccination with BNT162b2 started few weeks before vaccination with mRNA-1273.**

Figures 1-2 show the process for identifying infections in these cohorts, and Table 1 presents their demographic characteristics. Using the Kaplan–Meier estimator^14^, cumulative infection incidence among BNT162b2-vaccinated persons, with and without prior infection, was estimated at 0.14% (95% CI: 0.11-0.19%) and 0.93% (95% CI: 0.83-1.04%), respectively, after 63 days of follow-up (Figure 1). Incidence rates of infection were estimated, respectively, at 1.66 (95% CI: 1.26-2.18) and 11.02 (95% CI: 9.90-12.26) per 10,000 person-weeks. The incidence rate ratio was estimated at 0.15 (95% CI: 0.11-0.20).

**Table 1.** Demographic characteristics of matched cohorts that received the BNT162b2 and mRNA-1273 vaccines.

| Characteristics | Vaccination with the BNT162b2 vaccine |  |  | Vaccination with the mRNA-1273 vaccine |  |  |
| --- | --- | --- | --- | --- | --- | --- |
|  | Individuals with a prior PCR-confirmed infection | Individuals with no prior PCR-confirmed infection | p-value | Individuals with a prior PCR-confirmed infection | Individuals with no prior PCR-confirmed infection | p-value |
| **Median age (IQR) — years** | 39 (32-48) | 39 (32-48) | 0.972 | 40 (33-47) | 40 (33-47) | 0.869 |
| **Age group — no. (%)** |  |  |  |  |  |  |
| <20 years | 1,573 (3.1) | 1,573 (3.1) | 1.000 | 194 (0.8) | 194 (0.8) | 1.000 |
| 20-29 years | 7,282 (14.1) | 7,282 (14.1) |  | 3,481 (14.5) | 3,481 (14.5) |  |
| 30-39 years | 18,027 (35.0) | 18,027 (35.0) |  | 8,216 (34.2) | 8,216 (34.2) |  |
| 40-49 years | 13,593 (26.4) | 13,593 (26.4) |  | 7,972 (33.1) | 7,972 (33.1) |  |
| 50-59 years | 7,468 (14.5) | 7,468 (14.5) |  | 3,368 (14.0) | 3,368 (14.0) |  |
| 60-69 years | 2,830 (5.5) | 2,830 (5.5) |  | 704 (2.9) | 704 (2.9) |  |
| 70+ years | 713 (1.4) | 713 (1.4) |  | 117 (0.5) | 117 (0.5) |  |
| **Sex** |  |  |  |  |  |  |
| Male | 36,970 (71.8) | 36,970 (71.8) | 1.000 | 18,697 (77.7) | 18,697 (77.7) | 1.000 |
| Female | 14,516 (28.2) | 14,516 (28.2) |  | 5,355 (22.3) | 5,355 (22.3) |  |
| **Nationality<sup>†</sup>** |  |  |  |  |  |  |
| Bangladeshi | 3,728 (7.2) | 3,728 (7.2) | 1.000 | 2,066 (8.6) | 2,066 (8.6) | 1.000 |
| Egyptian | 3,470 (6.7) | 3,470 (6.7) |  | 1,748 (7.3) | 1,748 (7.3) |  |
| Filipino | 4,792 (9.3) | 4,792 (9.3) |  | 2,435 (10.1) | 2,435 (10.1) |  |
| Indian | 13,033 (25.3) | 13,033 (25.3) |  | 8,180 (34.0) | 8,180 (34.0) |  |
| Nepalese | 4,570 (8.9) | 4,570 (8.9) |  | 2,730 (11.4) | 2,730 (11.4) |  |
| Pakistani | 1,892 (3.7) | 1,892 (3.7) |  | 1,105 (4.6) | 1,105 (4.6) |  |
| Qatari | 9,700 (18.8) | 9,700 (18.8) |  | 1,047 (4.4) | 1,047 (4.4) |  |
| Sri Lankan | 1,490 (2.9) | 1,490 (2.9) |  | 1,114 (4.6) | 1,114 (4.6) |  |
| Sudanese | 1,259 (2.5) | 1,259 (2.5) |  | 481 (2.0) | 481 (2.0) |  |
| Other nationalities | 7,552 (14.7)<sup>‡</sup> | 7,552 (14.7)<sup>‡</sup> |  | 3,146 (13.1) | 3,146 (13.1) |  |

\*Cohorts were matched in a 1:1 ratio by sex, 5-year age group, nationality, and calendar week of first vaccine dose.
<sup>†</sup>Nationalities were chosen to represent the most numerous groups in the population of Qatar.
<sup>‡</sup> Individuals who received the BNT162b2 vaccine in Qatar comprised 96 other nationalities, while those who received the mRNA-1273 vaccine represented 78 other nationalities.

Cumulative infection incidence among mRNA-1273-vaccinated persons, with and without prior infection, was estimated at 0.06% (95% CI: 0.03-0.12%) and 0.08% (95% CI: 0.04-0.15%), respectively, after 63 days of follow-up (Figure 1). Incidence rates were estimated, respectively, at 1.55 (95% CI: 0.86-2.80) and 1.83 (95% CI: 1.07-3.16) per 10,000 person-weeks. The incidence rate ratio was estimated at 0.85 (95% CI: 0.34-2.05).

Infection incidence was low in these cohorts during a time of intense incidence in Qatar^6,7,15^, indicating that both vaccines were highly effective against the Alpha and Beta variants^6,7^, which dominated incidence^9^ (Methods). Still, prior infection of those BNT162b2-vaccinated further enhanced protection and reduced the incidence rate by 85% (6.6-fold) compared to those without prior infection. No evidence for such an effect was found for those mRNA-1273-vaccinated.

These findings are perhaps explained by the observed differences in effectiveness of these two vaccines against the Alpha and Beta variants, estimated in Qatar at 89.5% (95% CI: 85.9-92.3%) and 75.0% (95% CI: 70.5-78.9%) for BNT162b2, respectively^6^, and at 100% (95% CI: 91.8-100.0%) and 96.4% (95% CI: 91.9-98.7%) for mRNA-1273, respectively^7^.

The differences in effectiveness could have risen for a variety of reasons, such as differences in dosing, interval between doses, or the biology of both vaccines and their mechanisms of action. The dose of each of these two vaccines differed—it was 30-μg per dose for BNT162b2^4^ and 100 μg per dose for mRNA-1273^5^. This may have resulted in a more activated immune response for the mRNA-1273 vaccine than the BNT162b2 vaccine, and made the existence of prior immunity due to natural infection of no additional benefit for the mRNA-1273 vaccine. The interval between doses also differed and was one week longer for mRNA-1273^5^. Evidence suggests that a longer dose interval could be associated with improved protection after receiving the second dose^16^.

Limitations include identifying prior infection based on a record of a PCR-positive result, thereby missing those who may have been infected, but were unaware of their infection, or who did not seek testing by PCR to document the infection. Misclassification of prior infection status could lead to underestimation of the effect size of prior infection on vaccine protection. Depletion of the cohorts with prior infection due to COVID-19 mortality at time of the prior infection may have biased these cohorts toward healthier individuals with stronger immune responses. However, COVID-19 mortality has been low in Qatar’s predominantly young and working-age population^12,17^, and no evidence for such bias was found in the mRNA-1273 vaccine results, where the incidence rate was similar for those with and without prior infection.

We assessed risk of only documented infections, but other infections may have occurred and gone undocumented, perhaps because of minimal/mild or no symptoms. Our cohorts predominantly included working-age adults; therefore, results may not necessarily be generalizable to other population groups, such as children or the elderly. Matching was done for age, sex, nationality, and calendar week of the first vaccine dose, and could not be done for other factors, such as comorbidities or additional socio-demographic factors, as these were not available to study investigators. However, matching by age and sex may have served as a proxy given that co-morbidities are associated with older age and may be different between women and men. Matching by nationality may have also captured some of the occupational risk given the distribution of the labor force in Qatar^18-20^.

Imperfect assay sensitivity and specificity of PCR or antibody testing could have affected current or prior infection ascertainment. However, all PCR and serological testing was performed with extensively used, investigated, and validated commercial platforms with essentially 100% sensitivity and specificity (Methods). Unlike blinded, randomized clinical trials, the investigated observational cohorts were neither blinded nor randomized.

Our results demonstrate low infection incidence among those vaccinated with BNT162b2 or mRNA-1273, but among those vaccinated with BNT162b2, protection against infection was further enhanced and infection incidence was further reduced by prior infection. In contrast, those vaccinated with mRNA-1273 were as well protected as those who received the vaccine after a prior infection. These findings may have implications for the potential need of a booster vaccination.

## Data Availability

The dataset of this study is a property of the Qatar Ministry of Public Health that was provided to the researchers through a restricted-access agreement that prevents sharing the dataset with a third party or publicly. Future access to this dataset can be considered through a direct application for data access to Her Excellency the Minister of Public Health (https://www.moph.gov.qa/english/Pages/default.aspx). Aggregate data are available within the manuscript and its Supplementary information.

## Acknowledgements

We acknowledge the many dedicated individuals at Hamad Medical Corporation, the Ministry of Public Health, the Primary Health Care Corporation, and the Qatar Biobank for their diligent efforts and contributions to make this study possible. The authors are grateful for support from the Biomedical Research Program, the Biostatistics, Epidemiology, and Biomathematics Research Core, and the Genomics Core, all at Weill Cornell Medicine-Qatar, as well as for support provided by the Ministry of Public Health and Hamad Medical Corporation. The authors are also grateful for the Qatar Genome Programme for supporting the viral genome sequencing. The funders of the study had no role in study design, data collection, data analysis, data interpretation, or writing of the article. Statements made herein are solely the responsibility of the authors.

## Author contributions

LJA conceived and co-designed the study, led the statistical analyses, and co-wrote the first draft of the article. HC co-designed the study, performed the statistical analyses, and co-wrote the first draft of the article. All authors contributed to data collection and acquisition, database development, discussion and interpretation of the results, and to the writing of the manuscript.

All authors have read and approved the final manuscript.

## Competing interests

Dr. Butt has received institutional grant funding from Gilead Sciences unrelated to the work presented in this paper. Otherwise, we declare no competing interests.

## Methods

### Data sources and study design

Analyses were conducted using the centralized, integrated, and standardized national severe acute respiratory syndrome coronavirus 2 (SARS-CoV-2) databases compiled at Hamad Medical Corporation (HMC), the main public healthcare provider and the nationally designated provider for all Coronavirus Disease 2019 (COVID-19) healthcare needs. Through a nation-wide digital health information platform, these databases have captured all SARS-CoV-2-related data along with related-demographic details with no missing information since the start of the epidemic, including all records of polymerase chain reaction (PCR) testing, antibody testing, COVID-19 hospitalizations, vaccinations, infection severity classification per World Health Organization (WHO) guidelines^21^ (performed by trained medical personnel through individual chart reviews), and COVID-19 deaths, also assessed per WHO guidelines^22^. Every PCR test conducted in Qatar, regardless of location (outpatient clinic, drive-thru, or hospital, etc.), is classified on the basis of symptoms and the reason for testing (clinical symptoms, contact tracing, random testing campaigns (surveys), individual requests, routine healthcare testing, pre-travel, and port of entry). Qatar has unique demographics by sex and nationality, since expatriates from over 150 countries comprise 89% of the population^12,23^.

The nature of circulating SARS-CoV-2 virus was informed by weekly rounds of viral genome sequencing and multiplex, quantitative, reverse-transcription PCR (RT-qPCR) variant screening^24^ of randomly collected clinical samples^6,7,9-11^, as well as by the results of deep sequencing of wastewater samples^9^. The weekly rounds of viral genome sequencing from January 1-May 19, 2021 identified Beta (n=623; 50.9%), Alpha (n=193; 15.8%), Delta (n=43; 3.5%), and wild-type/undetermined variants (n=366; 29.9%) in 1,225 randomly collected, PCR-positive specimens^9,10^. Meanwhile, the weekly rounds of multiplex RT-qPCR variant screening from March 23-May 10, 2021 identified Beta-like (n=2,605; 66.4%), Alpha-like (n=970; 24.7%), and “other” variants (n=349; 8.9%) in 3,924 randomly collected PCR-positive specimens^9,11^. Sanger sequencing of the receptor binding domain of SARS-CoV-2 spike protein on 109 “other” specimens confirmed that 103 were Delta-like, 3 were B.1-like, and 3 were undetermined^9,11^.

All records of PCR testing in Qatar were examined in this study. Every individual that met the inclusion criteria in the national database, that is being vaccinated with BNT162b2 or mRNA-1273 and completing ≥14 days after the second vaccine dose, for each of these cohort studies, was classified based on infection status (with or without PCR-positive swab before the start of the study). Individuals were matched based on infection status on a 1:1 ratio by sex, 5-year age group, nationality (>75 nationality groups), and calendar week of first vaccine dose to control for differences in exposure risk^12,13^ and variant exposure^6,7,9-11^. Only matched samples were included in the analysis.

Further background on Qatar’s epidemic, such as on reinfections^25,26^, national seroprevalence surveys^12,18-20^, PCR surveys^12^, and other epidemiological studies can be found in previous publications on this epidemic^6,7,12,13,27-34^.

### Laboratory methods

Nasopharyngeal and/or oropharyngeal swabs (Huachenyang Technology, China) were collected for PCR testing and placed in Universal Transport Medium (UTM). Aliquots of UTM were: extracted on a QIAsymphony platform (QIAGEN, USA) and tested with real-time reverse-transcription PCR (RT-qPCR) using TaqPath™ COVID-19 Combo Kits (100% sensitivity and specificity^35^; Thermo Fisher Scientific, USA) on an ABI 7500 FAST (ThermoFisher, USA); extracted using a custom protocol^36^ on a Hamilton Microlab STAR (Hamilton, USA) and tested using AccuPower SARS-CoV-2 Real-Time RT-PCR Kits (100% sensitivity and specificity^37^; Bioneer, Korea) on an ABI 7500 FAST; or loaded directly into a Roche cobas® 6800 system and assayed with a cobas® SARS-CoV-2 Test (95% sensitivity, 100% specificity^38^; Roche, Switzerland). The first assay targets the viral S, N, and ORF1ab regions. The second targets the viral RdRp and E-gene regions, and the third targets the ORF1ab and E-gene regions. Antibodies against SARS-CoV-2 in serological samples were detected using a Roche Elecsys^®^ Anti-SARS-CoV-2 assay (99.5% sensitivity^39^, 99.8% specificity^39,40^; Roche, Switzerland), an electrochemiluminescence immunoassay that uses a recombinant protein representing the nucleocapsid (N) antigen for antibody binding. Results were interpreted according to the manufacturer’s instructions (reactive: optical density (proxy for antibody titer^41^) cutoff index ≥1.0 vs. non-reactive: optical density cutoff index <1.0).

All PCR tests were conducted at the Hamad Medical Corporation Central Laboratory or Sidra Medicine Laboratory, following standardized protocols.

### Statistical analysis

Descriptive statistics (frequency distributions and measures of central tendency) were used to characterize study samples. Significant associations were determined using two-sided p-values. The Kaplan–Meier estimator method^14^ was used to estimate the cumulative risk of documented infection. Cumulative risk was defined as the proportion of individuals identified with an infection during the study period among all eligible individuals in each cohort.

Incidence rates of documented infection in each cohort were calculated by dividing the number of infection cases identified during the study by the number of person-weeks contributed by all eligible individuals in the cohort. Incidence rates and corresponding 95% CIs were estimated using a Poisson log-likelihood regression model with the STATA 17.0^42^ *stptime* command. Follow-up person-time was calculated from the day each person completed 14 days after the second vaccine dose up to the infection swab, all-cause death, or end-of-study censoring (June 6, 2021). The incidence rate ratio and corresponding 95% CI were calculated using the exact method.

Statistical analyses were conducted in STATA/SE version 17.0^42^.

## Ethical approvals

The study was approved by the Hamad Medical Corporation and Weill Cornell Medicine-Qatar Institutional Review Boards with waiver of informed consent.

## Supplementary Material

**Supplementary Table 1.** STROBE checklist for cohort studies.

|  | Item No | Recommendation | Main Text page no |
| --- | --- | --- | --- |
| **Title and abstract** | 1 | (a) Indicate the study's design with a commonly used term in the title or the abstract<br>(b) Provide in the abstract an informative and balanced summary of what was done and what was found | 1<br>2 |
| **Introduction** |  |  |  |
| Background/rationale | 2 | Explain the scientific background and rationale for the investigation being reported | 3 |
| Objectives | 3 | State specific objectives, including any prespecified hypotheses | 3 |
| **Methods** |  |  |  |
| Study design | 4 | Present key elements of study design early in the paper | 3 |
| Setting | 5 | Describe the setting, locations, and relevant dates, including periods of recruitment, exposure, follow-up, and data collection | 15-18 |
| Participants | 6 | (a) Give the eligibility criteria, and the sources and methods of selection of participants. Describe methods of follow-up<br>(b) For matched studies, give matching criteria and number of exposed and unexposed | 15-17<br>16 |
| Variables | 7 | Clearly define all outcomes, exposures, predictors, potential confounders, and effect modifiers. Give diagnostic criteria, if applicable | 16 |
| Data sources/<br>measurement | 8* | For each variable of interest, give sources of data and details of methods of assessment (measurement). Describe comparability of assessment methods if there is more than one group | 15-16 |
| Bias | 9 | Describe any efforts to address potential sources of bias | 16 |
| Study size | 10 | Explain how the study size was arrived at | 16 & Figures 1-2 |
| Quantitative variables | 11 | Explain how quantitative variables were handled in the analyses. If applicable, describe which groupings were chosen and why | 16-17 |
| Statistical methods | 12 | (a) Describe all statistical methods, including those used to control for confounding<br>(b) Describe any methods used to examine subgroups and interactions<br>(c) Explain how missing data were addressed<br>(d) If applicable, explain how loss to follow-up was addressed<br>(e) Describe any sensitivity analyses | 16-17<br>NA<br>NA, see p.15<br>NA<br>NA |
| **Results** |  |  |  |
| Participants | 13* | (a) Report numbers of individuals at each stage of study—eg numbers potentially eligible, examined for eligibility, confirmed eligible, included in the study, completing follow-up, and analysed<br>(b) Give reasons for non-participation at each stage<br>(c) Consider use of a flow diagram | Figures 1-2 |
| Descriptive data | 14 | (a) Give characteristics of study participants (eg demographic, clinical, social) and information on exposures and potential confounders<br>(b) Indicate number of participants with missing data for each variable of interest<br>(c) Summarise follow-up time (eg, average and total amount) | Table 1<br>NA, see p.15<br>Table 1 |
| Outcome data | 15 | Report numbers of outcome events or summary measures over time | 3-4, Figure 3, and Supplementary Figure 1 |
| Main results | 16 | (a) Give unadjusted estimates and, if applicable, confounder-adjusted estimates and their precision (eg, 95% confidence interval). Make clear which confounders were adjusted for and why they were included<br>(b) Report category boundaries when continuous variables were categorized<br>(c) If relevant, consider translating estimates of relative risk into absolute risk for a meaningful time period | 3-4, Figure 3, and Supplementary Figure 1<br>16<br>NA |
| Other analyses | 17 | Report other analyses done—eg analyses of subgroups and interactions, and sensitivity analyses | NA |
| **Discussion** |  |  |  |
| Key results | 18 | Summarise key results with reference to study objectives | 4-5 |
| Limitations | 19 | Discuss limitations of the study, taking into account sources of potential bias or imprecision. Discuss both direction and magnitude of any potential bias | 5 |
| Interpretation | 20 | Give a cautious overall interpretation of results considering objectives, limitations, multiplicity of analyses, results from similar studies, and other relevant evidence | 5-6 |
| Generalisability | 21 | Discuss the generalisability (external validity) of the study results | 5 |
| **Other information** |  |  |  |
| Funding | 22 | Give the source of funding and the role of the funders for the present study and, if applicable, for the original study on which the present article is based | Acknowledgements |

Abbreviations: NA: not applicable;

**Supplementary Figure 1.**
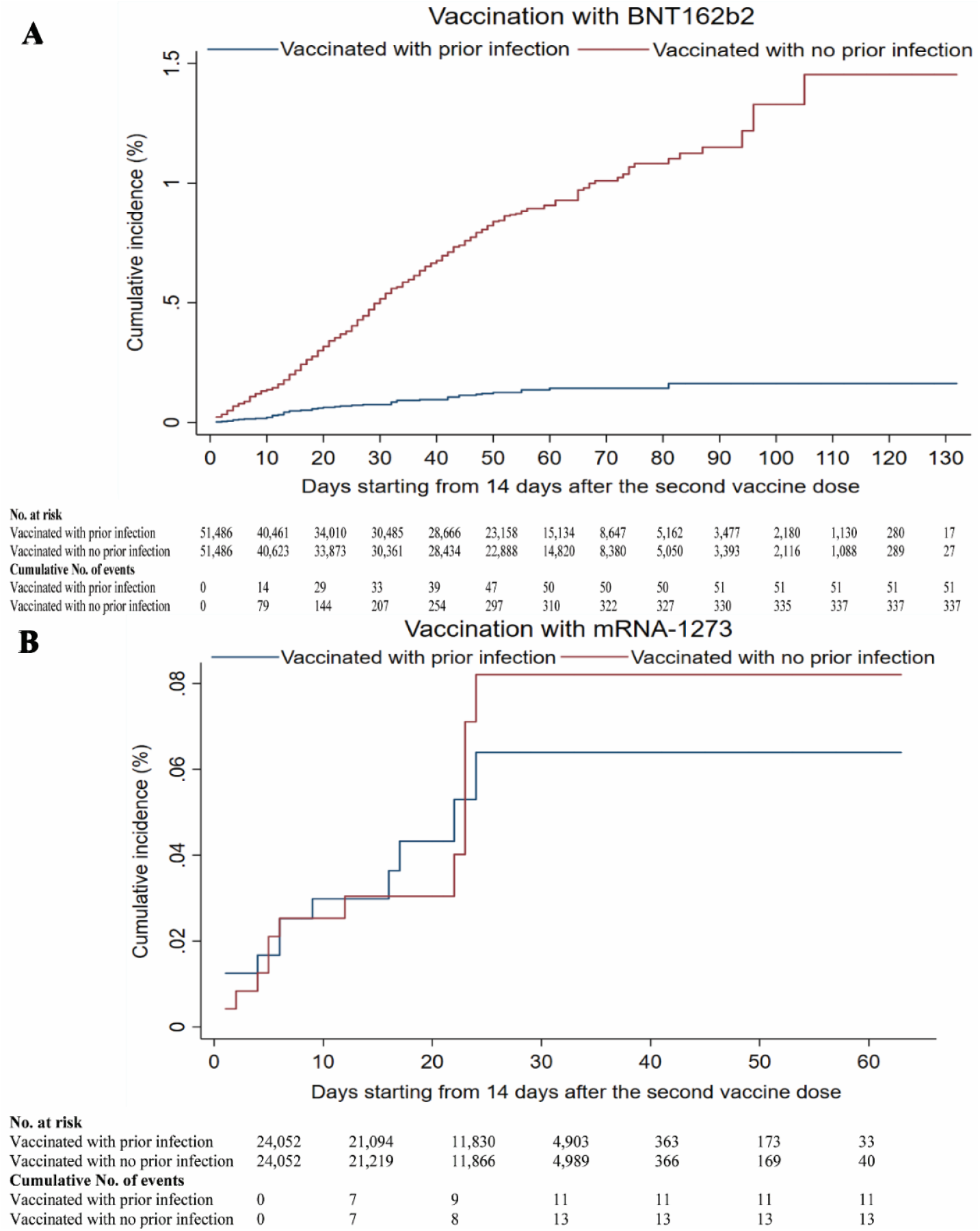
**Kaplan-Meier curves showing the cumulative incidence of documented SARS-CoV-2 infection in the national cohort of individuals who completed ≥14 days after the second vaccine dose and who had a prior PCR-confirmed infection, compared to the cumulative incidence of documented SARS-CoV-2 infection in the matched national cohort of individuals who completed ≥14 days after the second vaccine dose, but without prior PCR-confirmed infection. The curves compare vaccination with A) the BNT162b2 (Pfizer-BioNTech) vaccine and B) the mRNA-1273 vaccine. Cohorts were matched in a 1:1 ratio by sex, 5-year age group, nationality, and calendar week of the first vaccine dose. The cumulative infection incidence among the BNT162b2-vaccinated persons, with and without prior infection, was estimated at 0.16% (95% CI: 0.11-0.23%) and 1.45% (95% CI: 1.20-1.76%), respectively, after 132 days of follow-up.**

## Notes

### Competing Interest Statement

The authors have declared no competing interest.

